# Sub-district spatial heterogeneity in trachoma seroprevalence as populations approach elimination

**DOI:** 10.64898/2026.02.23.26346913

**Authors:** Ariktha Srivathsan, Everlyn Kamau, Ambahun Chernet, Getahun Ayenew, Tania A. Gonzalez, Eshetu Sata, Adisu Abebe, Zerihun Tadesse, E. Kelly Callahan, Karana Wickens, Sarah Gwyn, Diana L. Martin, Pearl Anne Ante-Testard, Jeremy D. Keenan, Thomas M. Lietman, Scott D. Nash, Benjamin F. Arnold

## Abstract

Programmatic decisions regarding surveillance and intervention for trachoma are made at the district level, reflecting an implicit assumption that transmission within districts is sufficiently homogeneous. However, as trachoma transmission declines, residual pockets of transmission may become spatially heterogeneous at sub-district scales. Using cluster-level data from 12 districts in Amhara, Ethiopia (2019–2023), we assess the spatial structure of Pgp3 antibody responses, a sensitive measure of infection transmission. Spatial clustering was evident in higher prevalence districts but diminished as populations approach elimination. We show that as districts approach elimination sub-district heterogeneity disappears, and district-level summaries are likely sufficient to guide decision-making.

## Background

Trachoma, caused by *Chlamydia trachomatis (Ct)*, remains the leading infectious cause of blindness worldwide^1^ although targeted public health interventions have led to substantial declines in prevalence across many endemic settings. Elimination as a public health problem is targeted by 2030.^2^ Nonetheless, persistent transmission continues in some regions, including Ethiopia.^3^ Programmatic decisions regarding surveillance and intervention are made at the district level^1^, reflecting an implicit assumption that transmission within these units is sufficiently homogeneous to justify aggregated decision-making.

Antibody responses, particularly prevalence of Immunoglobulin G (IgG) antibodies to the *Ct* antigen Pgp3 in young children, reflect past exposure and have emerged as valuable tools for monitoring transmission intensity^4^ and progress toward elimination.^5^ Model-based geostatistical approaches can further enhance monitoring through more precise prevalence estimation by borrowing strength across space, an advantage that may be especially important in low-transmission settings where subdistrict, cluster-level estimates are sparse and imprecise.^6,7^

Previous studies have demonstrated substantial sub-district heterogeneity in trachomatous inflammation—follicular (TF), a clinical sign of active inflammatory disease.^8,9^ Additionally, relationships between TF, *Ct* infection and serological markers can diverge, especially in low-transmission settings after repeated treatment,^10^ potentially due to different outcomes capturing transmission processes operating over distinct temporal and epidemiological scales. While recent work has quantified between-cluster heterogeneity in seroprevalence within standard survey designs,^11^ the extent to which seroprevalence exhibits meaningful spatial structure, and the implications of such structure for surveillance and programmatic decision-making, remain less well characterized.

As transmission declines, the remaining infections are hypothesized to become increasingly focal^12^, with elevated risk concentrated in geographically contiguous areas smaller than the district. If such spatial structure exists, district-level aggregation of prevalence may obscure epidemiologically and programmatically relevant heterogeneity. Conversely, if residual spatial dependence is weak at low prevalence, district-level summaries may remain sufficient. Here, we assess whether trachoma outcomes, particularly Pgp3 seroprevalence, exhibit meaningful sub-district spatial structure across a range of endemicity.

## Methods

### Study populations

We analyzed cluster-level trachoma surveillance data from the Global Trachoma Serology Data Repository^13^ collected in 12 districts in the Amhara region of Ethiopia between 2019 and 2023. Conjunctival swabs from children aged 1–5 years were tested for *Ct* infection by nucleic acid amplification using Polymerase Chain Reaction (PCR), and their dried blood spot (DBS) samples were tested for anti-Pgp3 antibodies.^3,14,15^ Children aged 1–9 years were examined for TF. Cluster sampling was used in each district survey, with villages as the primary sampling unit and households as the secondary sampling unit. Here, we estimated cluster-level prevalence for the three outcomes (TF, Pgp3 seroprevalence and *Ct* infection) separately by district. Further details of the surveys are in the **Supplementary Material**.

### Characterization of spatial heterogeneity

We assessed sub-district spatial autocorrelation in cluster-level prevalence using Moran’s I. For each district–outcome combination, Global Moran’s I was computed using k-nearest-neighbor spatial weights (k = 5). Neighbors were defined using Euclidean distance with row standardized spatial weights. Statistical inference was based on Monte Carlo permutation tests with 1000 simulations. Moran’s I estimates were treated descriptively with no adjustment for multiple testing.

### Spatial modeling of prevalence surfaces

To further characterize spatial structure, we fit spatial binomial models to cluster-level prevalence data within each district and outcome. Models were specified with the number of positive samples out of the total tested at each cluster as the outcome and a logit link. Spatial dependence was modeled using a Matérn covariance structure via a spatial random effect defined over cluster longitude and latitude and fit using maximum likelihood.

For district–outcome combinations with successfully fitted spatial models, we generated prediction surfaces by sampling ∼1,000 evenly spaced locations within each district polygon to visualize sub-district variation. All analyses were conducted in R version 4.5.1 and geographic coordinates were projected to UTM Zone 37N for spatial analyses.

## Results

### Study population and trachoma prevalence

The analysis included 12 districts with a range of trachoma endemicity across four studies conducted in Amhara. (**Figure 1A**). Eleven districts had 30 survey clusters each, while one urban district (Debre Birhan Town) had 26 clusters, for a total of 356 clusters that included 4,976 children aged 1–5 years and 8,399 aged 1–9 years. There was substantial variation in cluster-level prevalence, both between and within districts for all three outcomes (**Figure 1B**). In 1–5-year-olds, median cluster-level Pgp3 seroprevalence ranged from 0 to 22.2%, while *Ct* infection (PCR prevalence) was consistently lower, ranging from 0 to 4.8%, consistent with serology capturing cumulative exposure rather than current infection. TF in 1–9-year-olds ranged from 0% to 42.6% (**Supplementary Table 1**).

**Figure 1:**
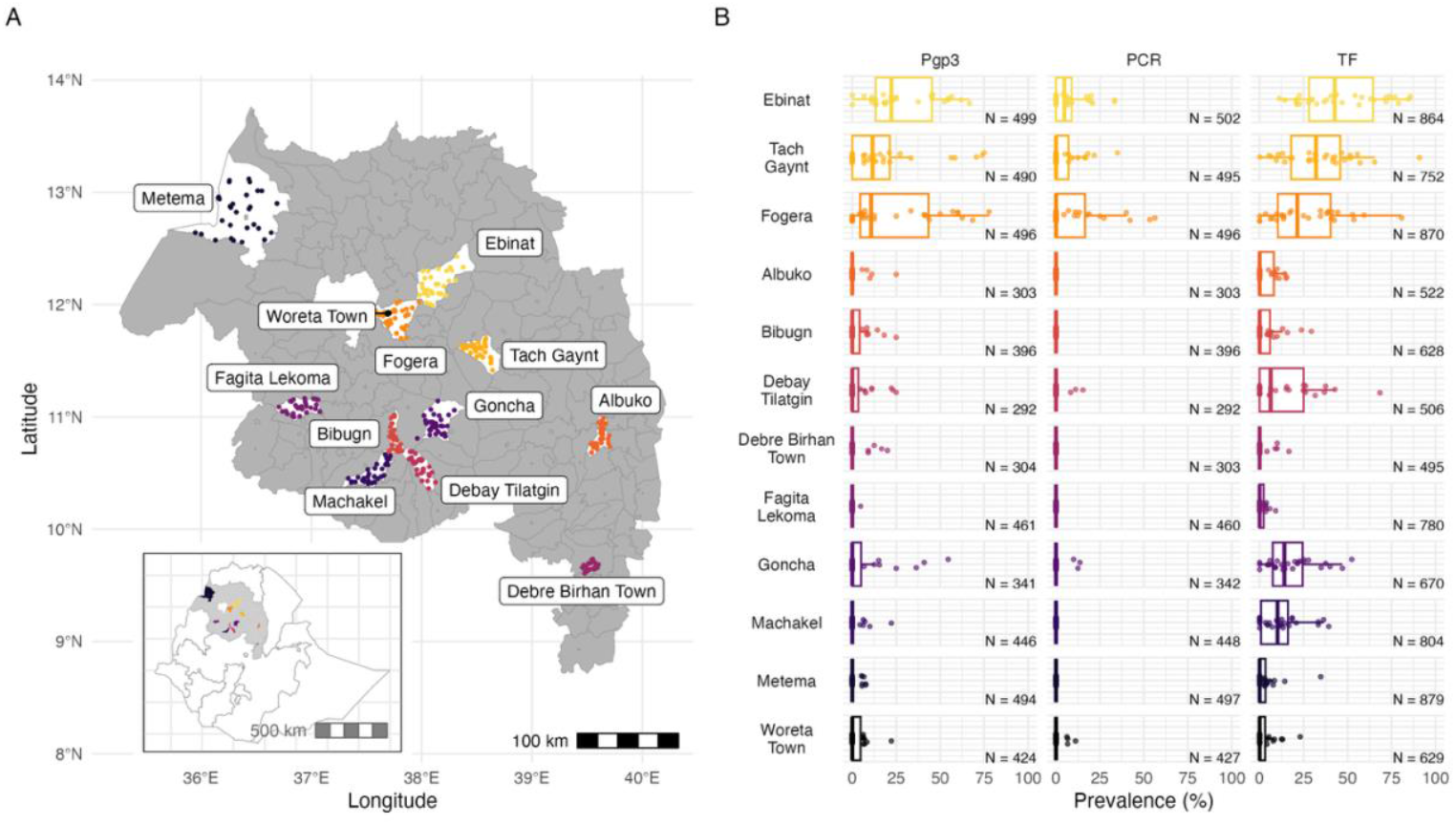
Study area and distribution of trachoma outcomes across districts in Amhara, Ethiopia. **A**. Map of the Amhara region with district boundaries, showing the locations of survey clusters included in the analysis (colored points). Districts included in subsequent analyses are highlighted in white. Inset map shows the position of Amhara Region within Ethiopia and the geographic extent of surveyed clusters. **B**. Distribution of cluster-level prevalence of three trachoma outcomes by district: IgG seroprevalence to *Chlamydia trachomatis* Pgp3 antigen in children ages 1-5 years, ocular *trachomatis* polymerase chain reaction (PCR) prevalence in children ages 1–5 years, and clinical trachomatous inflammation–follicular (TF) in children ages 1–9 years. Districts are ordered by median Pgp3 seroprevalence. Points indicate cluster-level prevalences and boxplots indicate the median and interquartile range within each district, illustrating substantial between-cluster heterogeneity. Point colors indicate rank of median cluster-level Pgp3 prevalence, ranging from yellow points in higher prevalence districts and dark blue indicating the lower prevalence districts. Sample sizes represent the total number of children tested for each outcome within each district.

### Sub-district spatial heterogeneity in Pgp3 seroprevalence

Evidence of sub-district spatial autocorrelation in Pgp3 seroprevalence varied systematically across the transmission gradient (**Figure 2, left panels**). Districts with higher Pgp3 seroprevalence exhibited moderate to strong positive spatial autocorrelation, including Ebinat (I = 0.36, p = 0.001), Tach Gaynt (I = 0.18, p = 0.022), and Fogera (I = 0.4, p = 0.001), consistent with spatial clustering at the sub-district level. In contrast, districts with lower Pgp3 seroprevalence showed weak or no evidence of spatial autocorrelation, including Woreta Town (I = 0.03, p = 0.23) and Metema (I = 0.06, p = 0.14). These results indicate that fine-scale spatial clustering of serological exposure is more apparent in districts with higher underlying transmission intensity and becomes less detectable as prevalence declines.

**Figure 2:**
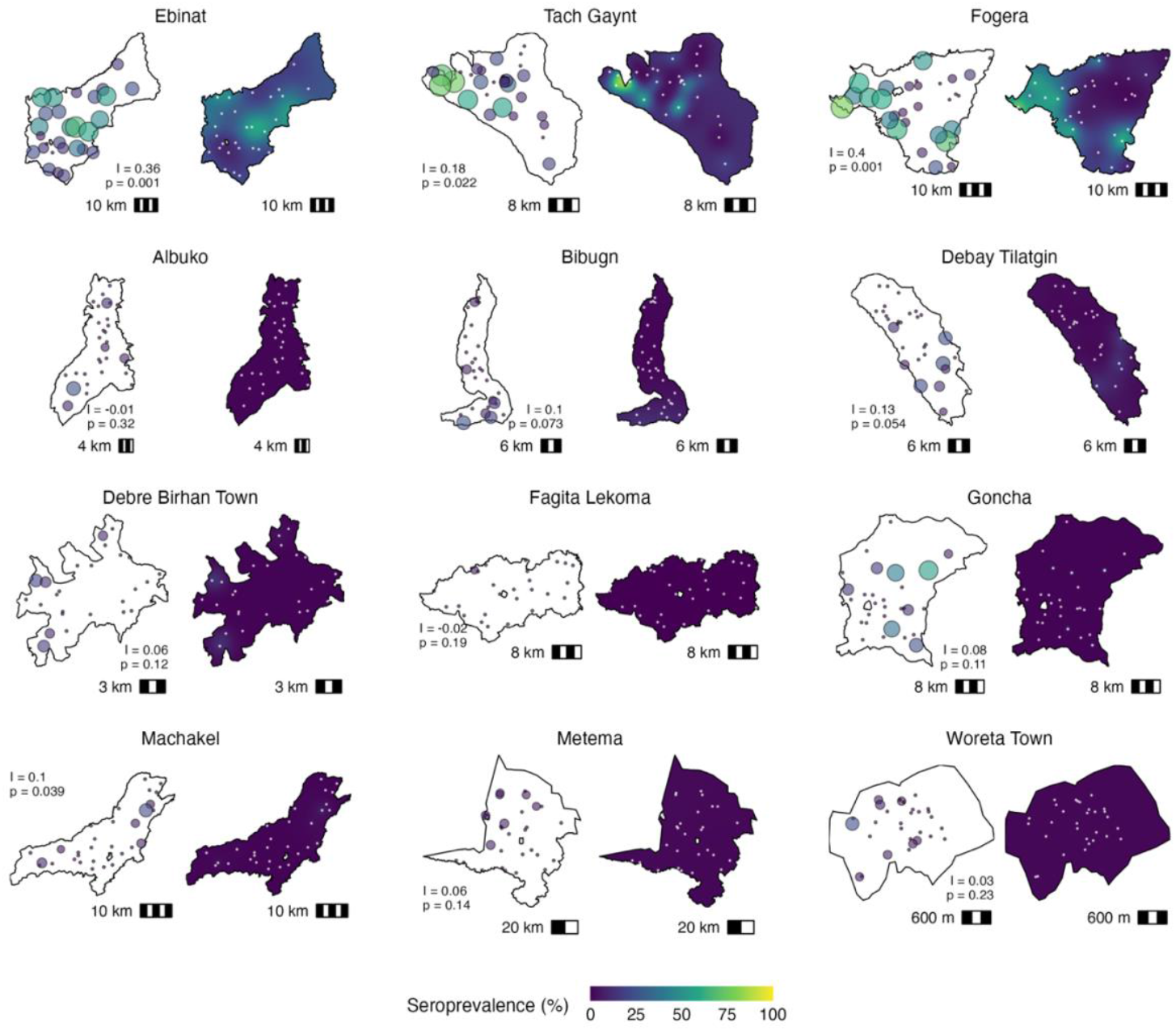
Observed and model-predicted spatial variation in Pgp3 seroprevalence by district. For each district, observed cluster-level Pgp3 seroprevalence is shown in the map on the <u>left</u> in each panel, with point size and color intensity proportional to prevalence at each survey cluster location. Moran’s I and p-value (Monte Carlo permutation test), quantifying the degree of spatial autocorrelation, are shown; values near zero indicate spatial randomness and positive values indicate spatial clustering. Maps on the <u>right</u> display model-predicted mean Pgp3 seroprevalence surfaces generated using district-specific geostatistical models. Models incorporate Matern spatial correlation functions to account for spatial dependence and to interpolate seroprevalence across unsampled locations within each district. White dots on the smoothed prediction surfaces indicate the locations of survey clusters used to fit each district-specific model. Districts with higher overall Pgp3 seroprevalence appear to exhibit more pronounced local spatial heterogeneity, in both observed data and model-predicted surfaces. In contrast, lower-prevalence districts demonstrate more diffuse, homogeneous spatial patterns.

Spatial prediction surfaces derived from district-specific spatial models provided a complementary visualization of sub-district variation in Pgp3 seroprevalence (**Figure 2, right panels**). In districts with significant positive spatial autocorrelation, predicted surfaces show spatially coherent areas, with elevated seroprevalence spanning multiple neighboring clusters, indicating that model-based smoothing captured structured variation beyond local cluster-level variation. In contrast, in districts with weak or non-significant Moran’s I estimates, prediction surfaces were largely flat, suggesting limited spatial structure despite observable between-cluster variation. Together, these results indicate that the utility of spatial modeling depends on the presence of detectable spatial dependence rather than on cluster-level heterogeneity alone.

In contrast to the smooth, spatially coherent patterns observed for Pgp3 seroprevalence, TF exhibited more fragmented spatial structure with substantial heterogeneity even in low-to-moderate prevalence districts, while PCR prevalence showed minimal spatial structure across most districts, with most clusters having zero prevalence and only isolated clusters with detectable infection, although one district (Tach Gaynt) had a statistically significant Moran’s I estimate (**Supplementary Figures 1, 2**).

## Discussion

Detectable sub-district spatial structure in Pgp3 seroprevalence varied systematically with underlying transmission intensity in Amhara. Districts with higher seroprevalence exhibited moderate to strong spatial autocorrelation, which weakened and became difficult to detect as seroprevalence declined and populations approached elimination.

Multiple plausible explanations may account for diminished spatial structure as populations approach elimination. First, incident infections may be increasingly sporadic as transmission declines, arising from stochastic importation events rather than sustained local transmission, and such exposures may be insufficient to induce and maintain detectable serologic responses. Second, sparse outcomes in low-prevalence settings reduce statistical power to detect spatial dependence even if focal structure exists. Third, historical exposure captured by serology may already be spatially homogenized in districts approaching elimination, particularly if recent declines in transmission have been geographically diffuse. This is consistent with the programmatic context: the lowest-prevalence districts in our analysis were undergoing surveillance surveys, having been classified as non-endemic for at least two years prior to data collection, suggesting that seroprevalence among children aged 1–5 years reflects recent rather than ongoing transmission.

The distinct spatial patterns observed for TF and *Ct* infection further support the interpretation that different trachoma markers capture transmission processes operating over different temporal and spatial scales. TF can persist for weeks to months following infection clearance, potentially explaining why fragmented clustering persists even as transmission declines (**Supplementary Figure 1**). In contrast, *Ct* infection represents a brief window of active infection, making it inherently sparse unless sampling coincides with active transmission. We thus observed minimal spatial structure of infection in all but one high-prevalence district (**Supplementary Figure 2**). Pgp3 serology, reflecting cumulative exposure, is therefore most likely to exhibit spatial structure in settings with sustained transmission, which diminishes as populations approach elimination.

From a programmatic perspective, these results have implications for both surveillance optimization and intervention targeting. Model-based geostatistical approaches have been shown to improve prevalence estimation^6,7^ and identify focal areas requiring intensified intervention^16^, and our findings suggest these methods are likely to be most informative for Pgp3 seroprevalence in moderate-to-higher transmission settings, where borrowing strength across space can enhance precision and reveal epidemiologically meaningful hotspots. However, in low-prevalence districts nearing elimination, the absence of detectable spatial structure in Pgp3 suggests that district-level summaries may remain appropriate for decision-making.

Important limitations include the exploratory nature of this analysis and restriction to a single region, though all indicator data are directly comparable across settings due to standardized measurement protocols. Future work needs to assess whether spatial autocorrelation in contiguous districts reveals transmission hotspots that span evaluation unit boundaries, particularly in settings where administrative borders may not align with underlying epidemiological connectivity. Our findings provide initial evidence that sub-district heterogeneity in Pgp3 seroprevalence is common in moderate-to-high transmission settings but disappears as populations approach elimination.

## Data Availability

Replication files are available through the Open Science Framework (https://osf.io/tbfhm). Geo-
located data is considered identifiable and so cannot be shared without appropriate IRB approval.

## Acknowledgements

The authors would like to thank Abbott for its donation of the m2000 RealTime molecular diagnostics system and consumables.

## Author Contributions

Following CRediT taxonomy: Conceptualization (SDN, BFA, AS); Data curation (EK, PAAT, DLM, SDN, BFA); Formal analysis (AS, BFA); Funding acquisition (SDN, BFA, TML); Investigation (AC, GA, TAG, ES, AA, ZT, EKC, KW, SG, DLM, SDN); Methodology (AS, BFA); Project administration (DLM, SDN, BFA); Software (AS, BFA); Supervision (BFA); Validation (AS, BFA); Visualization (AS, BFA); Writing – original draft (AS, EK, SDN, BFA); Writing – review & editing (all authors)

## Code and data availability

Replication files are available through the Open Science Framework (https://osf.io/tbfhm). Geo-located data is considered identifiable and so cannot be shared without appropriate IRB approval.

## Ethics statement

This study was approved by the Amhara Regional Health Bureau, the Federal Ministry of Science and Technology of Ethiopia, and by the Institutional Review Board (IRB) at Emory University (under protocol 079-2006). The protocols were further reviewed by Tropical Data staff (https://www.tropicaldata.org/) which ensures adherence to the WHO standardized approach for trachoma surveys. Informed verbal consent was obtained and recorded from all heads of households, study participants, and parents of participating children due to the extent of illiteracy in the region. Verbal assent was also obtained from participating minors in accordance with the Declaration of Helsinki. The secondary analysis protocol was reviewed and approved by the Institutional Review Board at the University of California, San Francisco (Protocol #20-33198).

## SUPPLEMENTARY MATERIAL

**Srivathsan et al**.

## Supplementary methods

### Study settings and trachoma surveys

#### Survey year 2019, Ebinat, Goncha, Debay Tilatgin, and Machakel districts^1^

Population-based cross-sectional surveys were conducted in 2019 in four districts in the Amhara region of Ethiopia: Ebinat, Goncha, Debay Tilatgin, and Machakel. These districts were all classified as <u>persistently endemic</u> for trachoma despite more than a decade of SAFE strategy implementation. Surveys used multi-stage cluster sampling with probability proportional to size to select communities, followed by random selection of household segments; all residents aged ≥1 year in selected households were eligible to participate. For the primary analytic population, children aged 1–9 years were examined for trachoma clinical signs by graders trained and certified in Tropical Data methods, were swabbed for *Chlamydia trachomatis* infection testing, and provided dried blood spot specimens for serological testing. Biospecimen were stored under cold-chain conditions until testing. Infection was measured by the Abbot Realtime assay on the m2000 PCR system at the Trachoma Molecular Laboratory at the Amhara Public Health Institute. Antibodies to *Chlamydia trachomatis* were measured using a multiplex bead assay targeting the Pgp3 antigen, a marker of cumulative exposure. Dried blood spots were eluted and assayed at the U.S. Centers for Disease Control and Prevention, with antibody responses quantified as median fluorescence intensity with background subtraction (MFI-BG). Seropositivity thresholds for Pgp3 were defined using receiver operating characteristic–based cutoffs derived from reference panels of known positive and negative sera. Age-specific *Chlamydia trachomatis* infection and seroprevalence provided complementary indicators to clinical trachoma signs for assessing fine-scale spatial heterogeneity in transmission in persistently endemic settings.

#### Survey year 2021, Metema and Woreta Town districts^2^

Population-based, multi-stage cluster surveys were conducted in 2021 in the trachoma-endemic districts of Metema and Woreta Town in the Amhara region, Ethiopia, a setting with historically high transmission that has undergone repeated rounds of the SAFE strategy and is now transitioning to <u>post-elimination surveillance</u>. Clusters (communities) were selected using probability proportional to size sampling, followed by random selection of household segments; all resident children were eligible for participation. The primary study population comprised children aged 1–9 years. Following informed consent, children were graded for trachoma clinical signs by graders trained and certified in Tropical Data methods, they were swabbed for *Chlamydia trachomatis* infection testing, and dried blood spots were collected via finger prick from children Biospecimen were stored under cold-chain conditions. Infection was measured by the Abbot Realtime assay on the m2000 PCR system at the Trachoma Molecular Laboratory at the Amhara Public Health Institute. Serological responses to *Chlamydia trachomatis* were measured using a multiplex bead assay targeting antibodies to the Pgp3 antigen, a well-established marker of cumulative exposure. Dried blood spots were eluted and assayed at the U.S. Centers for Disease Control and Prevention, with antibody responses quantified as median fluorescence intensity with background subtraction (MFI-BG). Seropositivity thresholds were defined using receiver operating characteristic–based cutoffs derived from reference panels of known positive and negative specimens. Age-specific seroprevalence were subsequently estimated to characterize recent and historical transmission intensity. Age-specific *Chlamydia trachomatis* infection and seroprevalence provided complementary indicators to clinical trachoma signs for assessing fine-scale spatial heterogeneity in transmission in low-prevalence, post-MDA settings.

#### Survey year 2022, Albuko, Bibugn, Debre Birhan town, and Fagita Lekoma districts^3^

Population-based, multi-stage cluster surveys were conducted in 2022 in Albuko, Bibugn, Debre Birhan town, and Fagita Lekoma, districts <u>close to or below the elimination</u> threshold of 5% TF. Clusters (communities) were selected using probability proportional to size sampling, followed by random selection of household segments; all resident children were eligible for participation. The primary study population comprised children aged 1–9 years. Following informed consent, children were graded for trachoma clinical signs by graders trained and certified in Tropical Data methods, they were swabbed for *Chlamydia trachomatis* infection testing, and dried blood spots were collected via finger prick from children. Biospecimen were stored under cold-chain conditions until testing. Infection was measured by the Abbot Realtime assay on the m2000 PCR system at the Trachoma Molecular Laboratory at the Amhara Public Health Institute. Serological responses to *Chlamydia trachomatis* were measured using a multiplex bead assay targeting antibodies to the Pgp3 antigen, a well-established marker of cumulative exposure. Dried blood spots were eluted and assayed at the U.S. Centers for Disease Control and Prevention, with antibody responses quantified as median fluorescence intensity with background subtraction (MFI-BG). Seropositivity thresholds were defined using receiver operating characteristic–based cutoffs derived from reference panels of known positive and negative specimens. Age-specific seroprevalence were subsequently estimated to characterize recent and historical transmission intensity. Age-specific *Chlamydia trachomatis* infection and seroprevalence provided complementary indicators to clinical trachoma signs for assessing fine-scale spatial heterogeneity in transmission in low-prevalence, post-MDA settings.

#### Survey year 2023, Fogera and Tach Gaynt districts

Population-based, multi-stage cluster surveys were conducted in 2023 in Fogera and Tach Gaynt districts, districts classified as <u>experiencing persistent trachoma</u>. <u>Similar to the earlier surveys, c</u>lusters (communities) were selected using probability proportional to size sampling, followed by random selection of household segments; all resident children were eligible for participation. The primary study population comprised children aged 1–9 years. Following informed consent, children were graded for trachoma clinical signs by graders trained and certified in Tropical Data methods, they were swabbed for *Chlamydia trachomatis* infection testing, and dried blood spots were collected via finger prick from children. Biospecimen were stored under cold-chain conditions until testing. Infection was measured by the Abbot Realtime assay on the m2000 PCR system at the Trachoma Molecular Laboratory at the Amhara Public Health Institute. Serological responses to *Chlamydia trachomatis* were measured using a multiplex bead assay targeting antibodies to the Pgp3 antigen, a well-established marker of cumulative exposure. Dried blood spots were eluted and assayed at the U.S. Centers for Disease Control and Prevention, with antibody responses quantified as median fluorescence intensity with background subtraction (MFI-BG). Seropositivity thresholds were defined using receiver operating characteristic–based cutoffs derived from reference panels of known positive and negative specimens. Age-specific seroprevalence and seroconversion rates were subsequently estimated to characterize recent and historical transmission intensity. Age-specific *Chlamydia trachomatis* infection and seroprevalence provided complementary indicators to clinical trachoma signs for assessing fine-scale spatial heterogeneity in transmission in persistently endemic settings.

## Supplementary Figures

**Supplementary Figure 1:**
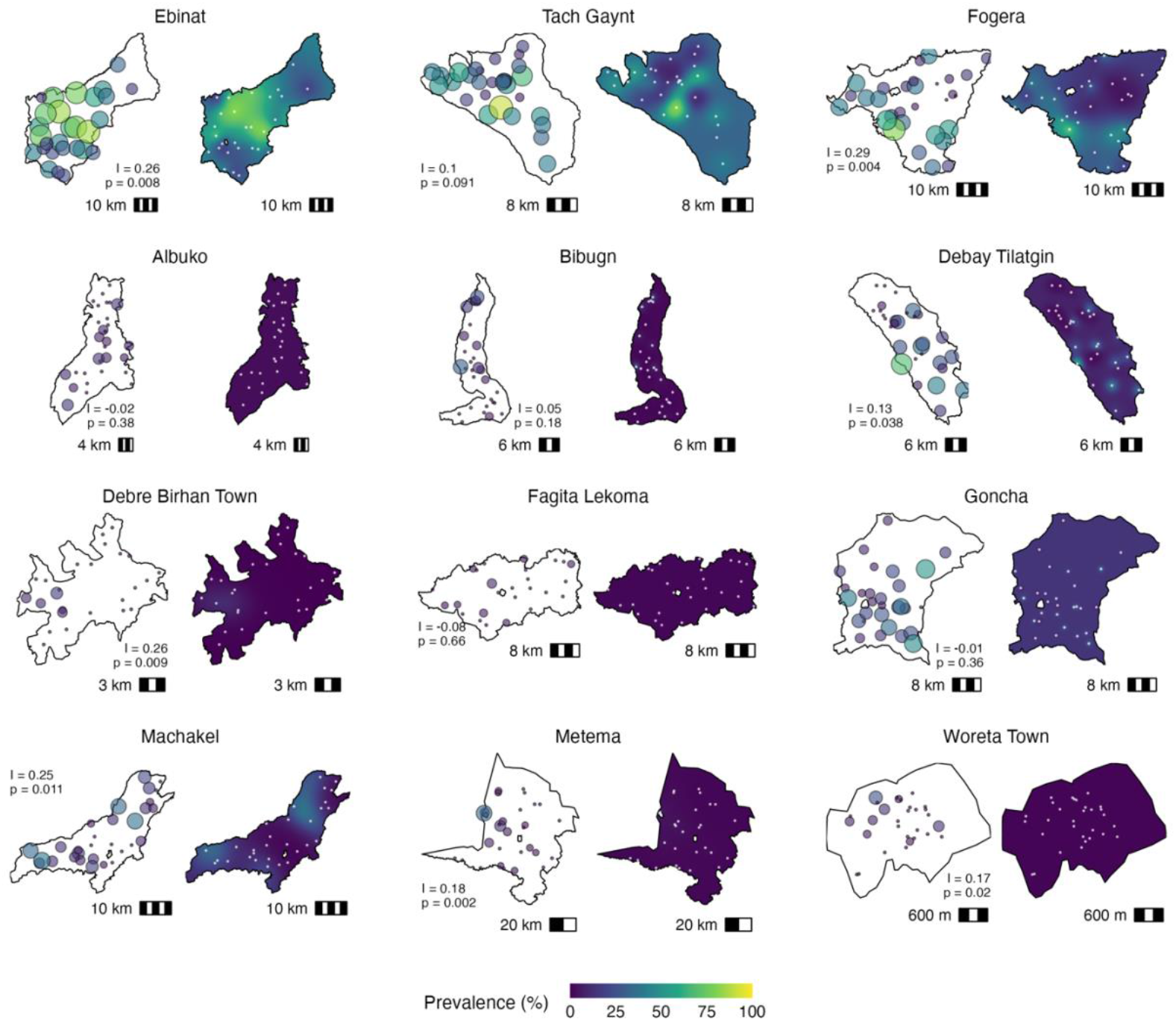
Observed and model-predicted sub-district spatial variation in trachomatous inflammation–follicular (TF) prevalence by district. For each district, observed cluster-level TF prevalence in children aged 1–9 years is shown in the left panel, with point size and color intensity proportional to prevalence at each survey cluster location. Right panels display model-predicted mean TF prevalence surfaces generated using district-specific geostatistical models. Models incorporate Matern spatial correlation functions to account for spatial dependence and to interpolate prevalence across unsampled locations within each district. White dots on the smoothed prediction surfaces indicate the locations of survey clusters used to fit each district-specific model. TF prevalence exhibits greater local variability than Pgp3, with substantial heterogeneity within districts, including districts where median prevalence is low or moderate. Compared with the smoother and more spatially coherent patterns observed for Pgp3 seroprevalence, TF displays more fragmented spatial structure, with more isolated elevated clusters rather than forming contiguous focal regions.

**Supplementary Figure 2:**
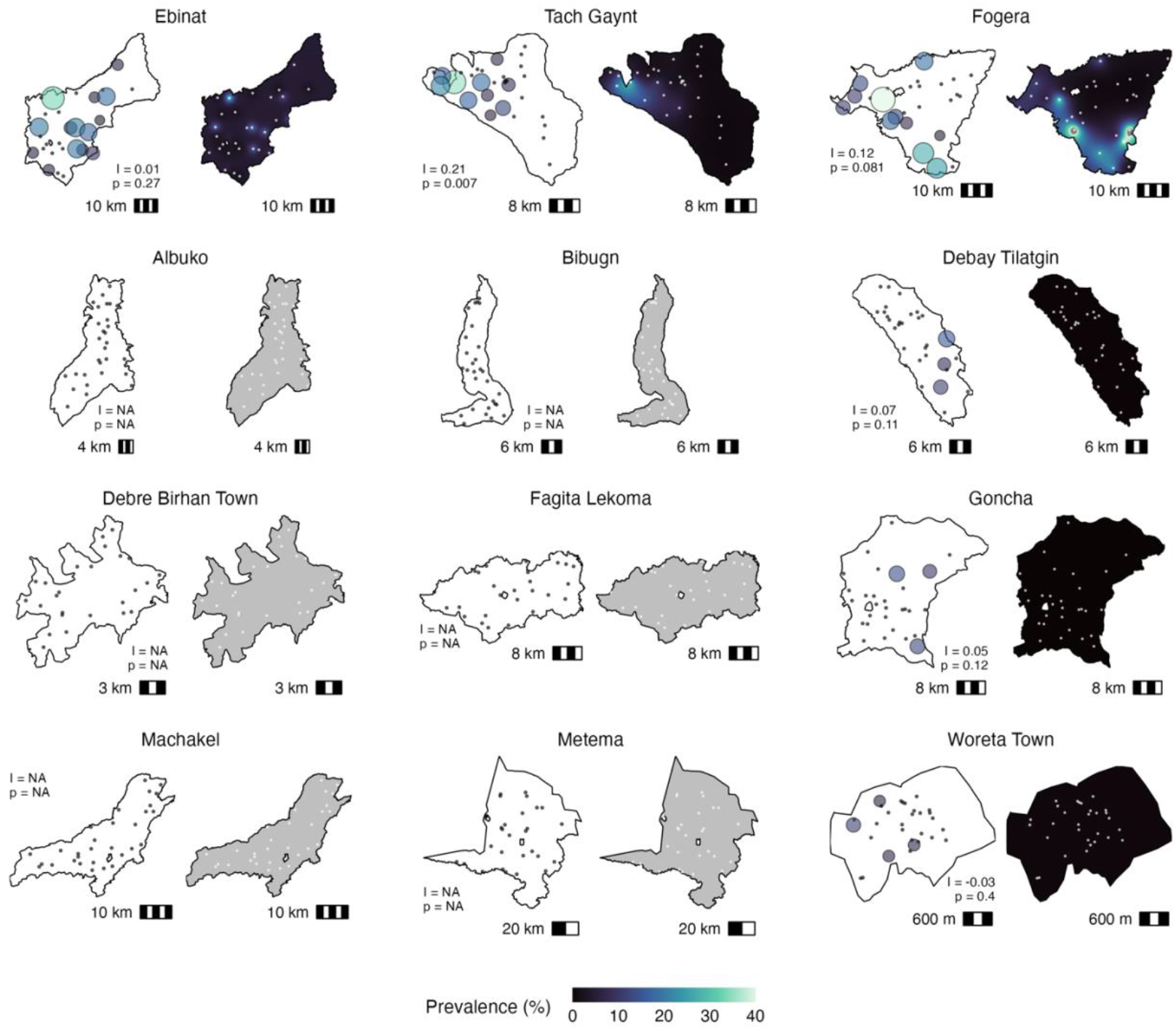
Observed and model-predicted sub-district spatial variation in *C. trachomatis* infection by district. For each district, observed cluster-level *Ct* infection, measured as PCR prevalence, in children aged 1–5 years is shown in the left panel, with point size and color intensity proportional to prevalence at each survey cluster location. Right panels display model-predicted mean PCR prevalence surfaces generated using district-specific geostatistical models. Models incorporate Matern spatial correlation functions to account for spatial dependence and to interpolate prevalence across unsampled locations within each district. White dots on the smoothed prediction surfaces indicate the locations of survey clusters used to fit each district-specific model. Grey surfaces indicate six districts where geostatistical models could not be fit due to uniformly low prevalence across all clusters, precluding estimation of spatial correlation parameters. Statistically significant spatial autocorrelation was observed only in Tach Gaynt (Moran’s I = 0.21; P = 0.007). In most other districts, including the highest-prevalence districts, multiple clusters had no *Ct* infection and detectable infections were observed in fairly isolated locations, showing minimal detectable spatial structure.

## Supplementary Table

**Supplementary Table 1:** Median and interquartile range of prevalence of three trachoma outcomes by district, displayed in Figure 1 of the main text. Pgp3 measures IgG seroprevalence among children ages 1-5-years-old. TF measures clinical signs of trachoma (trachomatous inflammation–follicular) among children ages 1-9-years-old. PCR measures ocular infection with *Chlamydia trachomatis* among children 1-5-years-old.

| District | Prevalence: Median (IQR) |  |  |
| --- | --- | --- | --- |
|  | Pgp3 | TF | PCR |
| Ebinat | 22.2 (31.9) | 42.6 (36.3) | 4.8 (8.9) |
| Tach Gaynt | 11.5 (21.3) | 32.1 (28) | 0 (7.1) |
| Fogera | 10.8 (38.8) | 21.4 (30) | 0 (16.6) |
| Albuko | 0 (0) | 0 (8) | 0 (0) |
| Bibugn | 0 (4.2) | 0 (5.9) | 0 (0) |
| Debay Tilatgin | 0 (3.4) | 6.3 (25) | 0 (0) |
| Debre Birhan Town | 0 (0) | 0 (0) | 0 (0) |
| Fagita Lekoma | 0 (0) | 0 (2.3) | 0 (0) |
| Goncha | 0 (5) | 14 (17) | 0 (0) |
| Machakel | 0 (0) | 10.2 (15.2) | 0 (0) |
| Metema | 0 (0) | 0 (3.3) | 0 (0) |
| Woreta Town | 0 (4.7) | 0 (3.1) | 0 (0) |

